# The Schema Method: Using Quantified Learning to Innovate, Augment, Assess and Analyze Learning in Medicine

**DOI:** 10.1101/2022.09.06.22279624

**Authors:** Deepu Sebin, Vishwin Doda, Skanthvelan Balami

**Author notes:** Corresponding Author: Dr. Vishwin Doda.

## Abstract

**Introduction:** The predominant method of learning Medicine at its core has remained unchanged for decades. This stagnancy creates a need for making learning more effective, insightful, and quantified. ‘Schema’ achieves this through e-learning, active feedback, and quantified learning by granulating the medical curriculum into specific subtopics selected based on the crucial knowledge that a competent medical learner must possess, hereafter referred to as ‘yield.’ This particular study aims to analyze medical students’ multidimensional competency in solving clinical scenario-based MCQs pertaining to vertically integrated topics derived from the ‘Schema.’

**Methods:** A retrospective study was conducted by analyzing the user data of a leading e-learning platform for medical students. For the purposes of this study, twenty such “high-yield” Schema topics were shortlisted as being the most crucial knowledge areas. Students’ performance in solving a fixed set of SOC-MCQs of these Schema topics was used to gauge their competence. Performance variation over five years (2018-2022) was also analyzed to study the changing patterns in topic-specific performance.

**Results:** A total of 20 Schema topics were shortlisted, consisting of 128 MCQs. The number of participants solving each Single Option Correct Multiple Choice Question (SOC-MCQ) ranged from 60,080 to 2,06,672. A significant improvement in the Net Delta was observed in 9 topics. Performance in other topics showed either no significant change or a significant downtrend.

**Conclusion:** A significant performance uptrend (ND = 128%) was observed in Anaphylaxis, Basic Lifesaving Skills, ST-Elevated Myocardial Infarction, Glasgow Coma Scale, and subdural hemorrhage & Syndromic management of Sexually Transmitted Infections, most of which are either acute or emergency conditions. A significant downtrend in performance was seen in Schema topics such as Asthma management, Hypertension management, Diabetic Ketoacidosis, and Subarachnoid hemorrhage pertaining to chronic conditions. Several hypotheses for these findings can be derived, the validities and collective impacts of which can be explored in more in-depth and broader studies in the future.

## Introduction & Background

The advent of technology in learning and teaching has brought about a new era in the field of education. (1) Furthermore, the enlightening changes in our understanding of how learning itself works have motivated the medical fraternity across the globe to explore techniques such as Self Directed Learning (SDL), Problem Based Learning (PBL), and Objective Assessment via Single Option Correct - Multiple Option Questions (SOC-MCQs) in the last few years. (2–5). Learning augmented with such methods has consistently been more effective than traditional methods. (6–10) However, one area that primarily lies unexplored is Quantified Learning, a method that can potentially change how we learn, assess, and measure competence in the field of Medicine.

*Quantified learning* is a novel learning method based on data cognition and aims to provide a personalized learning experience. It is a closed-loop system with adaptive feedback consisting of the data, learners, stakeholders, and the learning service itself. Z.Talib defines Quantified Learning as the “use of technology to help provide more granular and predictable education and education outcomes.” (11), (12).

The critical areas of Quantified learning are:

i. Breaking down the acquisition of a skill or knowledge area into small pieces
ii. Identifying and delivering knowledge courses for each of those areas
iii. Assessing and diagnosing learning and mastery
iv. Providing appropriate feedback

Some examples of quantified learning are self-directed learning, case-based learning, blended learning, active learning, and feedback-based learning. Quantified learning and related practices have been shown to impact students’ learning and motivation to learn positively. (12-15)

### The prevalent learning method

Medical learning has remained more or less the same through the decades. The current learning method largely consists of two domains:

i. Theoretical: The theoretical aspect mainly consists of textbook-based learning combined with peer-to-peer learning or learning via didactic lectures. The textbooks are segregated by subjects such as Pediatrics, Medicine and Surgery.. The standard curriculum of respective subjects is prescribed, usually assessed via written exams or oral vivas.
ii. Practical/Clinical Mainly involves intra-departmental clinical case discussions, patient interaction, clinical demonstrations, observation & assistance of surgery, and hands-on clinical training, either with simulation mannequins or live subjects. It is traditionally tested via clinical demos or Objective Structured Clinical Examinations (OSCE).

This subject-wise segregated approach has stood the test of time but is devoid of continuous feedback, suitable performance indicators, or quantified learning. Moreover, it leaves little room for intuitional vertical integration of knowledge points which is increasingly being acknowledged to be of utmost importance in the understanding & practice of Medicine. (16–19) As a result, the overall competency of a medical student or doctor in specific areas of medical knowledge is still considered an abstract and unquantifiable parameter rather than a tangible metric. A clear need to develop more objective and quantifiable methods to assess competence in medical practice has been felt, and various attempts have been made to the same end. (20–22)

### Scope for Improvement in Medical Learning

A broad consensus has existed for decades that calls for significant changes in the medical education system.(23,24). One primary goal in modern medical education systems is to ensure that learning & assessment takes place in a quantifiable manner and valuable feedback to the learner regarding their competency is actively accessible.(25)

The Carnegie report undertaken after 100 years of observance after the acclaimed Flexner report lists the following principles to be followed while reforming medical education:(26)

- It aims to have learners develop the motivation and skill to teach themselves.
- Medical education must ensure thorough assessment that learners achieve predetermined standards of competence with respect to knowledge and performance in core domains.
- Assessment must go beyond what learners know and can do to address learners’ ability to identify gaps and next steps for learning.

These principles are central to the systems of Quantified Self and self-directed learning. The quantified self is a person that uses technology to record and track information about a part of life in order to improve a skill. (27) Improved physician skills and, by extension, improved patient care are bound to be the outcome of such a dynamic & meticulous model of learning. Recognizing and defining such crucial competencies is a core challenge facing medical education today.(28)

Another challenge is presented by the amazing vastness of the medical curriculum itself. In addition, the definitive list of “crucial topics in medicine” is in itself a dynamic entity that is bound to keep changing with the ever-changing tides of healthcare challenges.(2) COVID-19, for example, has brought many medical topics previously considered “non-crucial” by medical academia and learning methods to the forefront of Medicine.(29–31) One possible solution that addresses these challenges is a well-curated, up-to-date, and interactive system rooted in Quantified Learning that promotes expertise in critical concepts, aids in the elimination of knowledge gaps, and provides the learner with invaluable feedback to allow self-improvement.

### Schema & The Schema Method

An innovative app-based e-learning solution was created to reorganize the standard medical curriculum into a collection of granular high-yield topics that not only accounts for their cruciality in the study & practice of Medicine but also seamlessly integrates them across each relevant subject. The same collective database of these high-yield granular sub-topics presented in an interactive feedback system is what we named Schema. At the time of completion of this study, 2048 topics had been identified & listed in Schema, with each Schema topic linked to multiple MCQs. Each of these MCQs is clinically relevant, standardized, peer-reviewed, and displays a solution with an in-depth explanation upon being solved. The explanation helps the learning & assessment parts proceed simultaneously, giving rise to a feedback loop that leads to consistent improvement in knowledge. Each MCQ is allotted a unique ID to aid performance tracking.

In this study, a tendency was noted to solve these MCQs while advancing through the prescribed medical course of the study or addressing knowledge gaps. Visual feedback was also provided to the learner that consisted of percentage correctness in each subtopic, the extent of coverage of the curriculum, and a color-coded change with red for low cumulative performance in a particular Schema and green for a Schema topic completed with a good score.

Since these Schema topics are high-yield and are essential to be known by medical students & practicing physicians alike, the quantified measurement of how adept the learner is in these topics, along with the extent of coverage of Schema topics & constant feedback to the user, has the potential to reflect the overall competency of the practitioner effectively. This self-directed, integrated, feedback-based method of learning these granulated topics by the quantified self is what we term “The Schema Method.”

## Aims & Objectives

### Aim

This study aims to leverage Schema to assess & quantify the performance of medical students in the most crucial sub-topics and study the change in performance over time with the help of novel metrics.

### Objectives

- To assess the current knowledge & performance of medical students in the aforementioned crucial Schema subtopics.
- To study the variation of performance of medical students in the crucial Schema subtopics
- To obtain a net change in the performance of crucial Schema topics over five years

## Materials & Methods

### Type of Study

Retrospective Analytical Study

### Study Population

Current users of a leading online medical education app that are one of the following:

- Medical students in First Year, Second Year, Third Year, Final Year MBBS
- Medical Interns
- MBBS graduates

### Sample Size

Each MCQ was solved voluntarily by each participant based on the need to either advance through the prescribed MBBS course, address an area of personally quantified knowledge gaps, or self-assess oneself in a particular subtopic. It is assumed that each MCQ was solved independently for all purposes of this study and the contributing analysis.

### Study Design

25 of the most crucial Schema topics that medical professionals must ideally be adept in were chosen by the panel of authors based on applicability, disease prevalence, or commonly encountered medical emergencies to quantify the change in performance of learners over time. Each subtopic is linked to many MCQs that test the core learning principles of each subtopic.

**Table 1:**
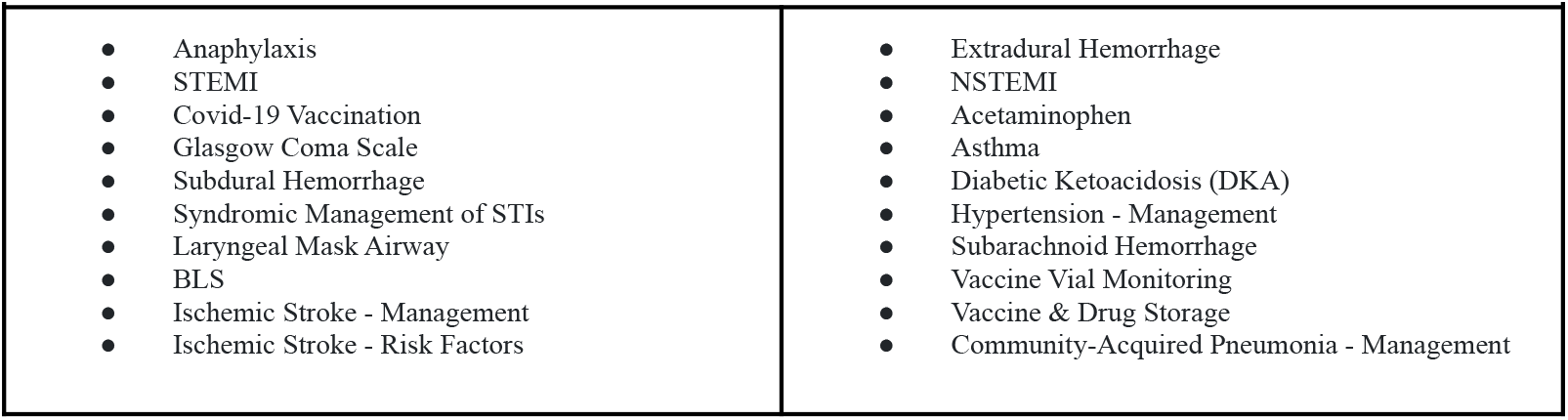
List of crucial Schema subtopics shortlisted for this study.

### Statistical Analysis

The cumulative data of all MCQs attempted during 2018-2022 was compiled and analyzed to gauge the performance of participants & its variation during the five year period. Google Sheets (Alphabet Inc.©) was used for statistical analysis. MCQs were allocated into their respective Schema with the help of a unique ID (n). For each MCQ, the following data parameters were collected:

- A total number of students that solved the MCQ (*T*_*n*_)
- Number of students that answered the MCQ correctly (*C*_*n*_)

The collected data was used to derive the following statistics:

1. ***Schema correctness percentage (SCP):*** The cumulative performance in a Schema subtopic was calculated for each year by taking the average correctness of all MCQs in a Schema topic. This metric allowed the deduction of variation of students’ performance in a topic over time.

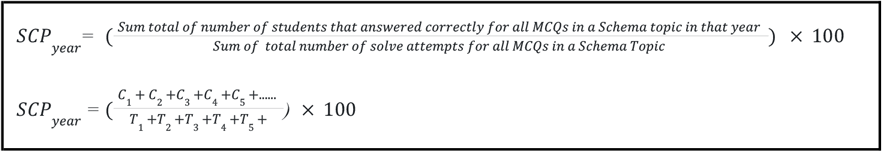
2. ***Delta:*** Represents the change in the percentage correctness of all the MCQs in a Schema topic compared to the last year. Delta for the year 2018 was considered to be zero since the data timeline of this study commenced in 2018. Since delta represents a change in the percentage correctness, it can be either a positive value (representing increased correctness as compared to the previous year) or a negative value (increased correctness as compared to the previous year.

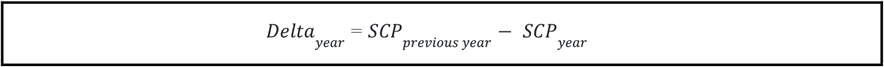
3. ***Net Delta (ND):*** Represents the net change in the percentage correctness of all the MCQs in a Schema topic over the course of 5 years.

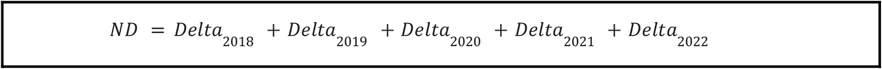

**Figure 1:**
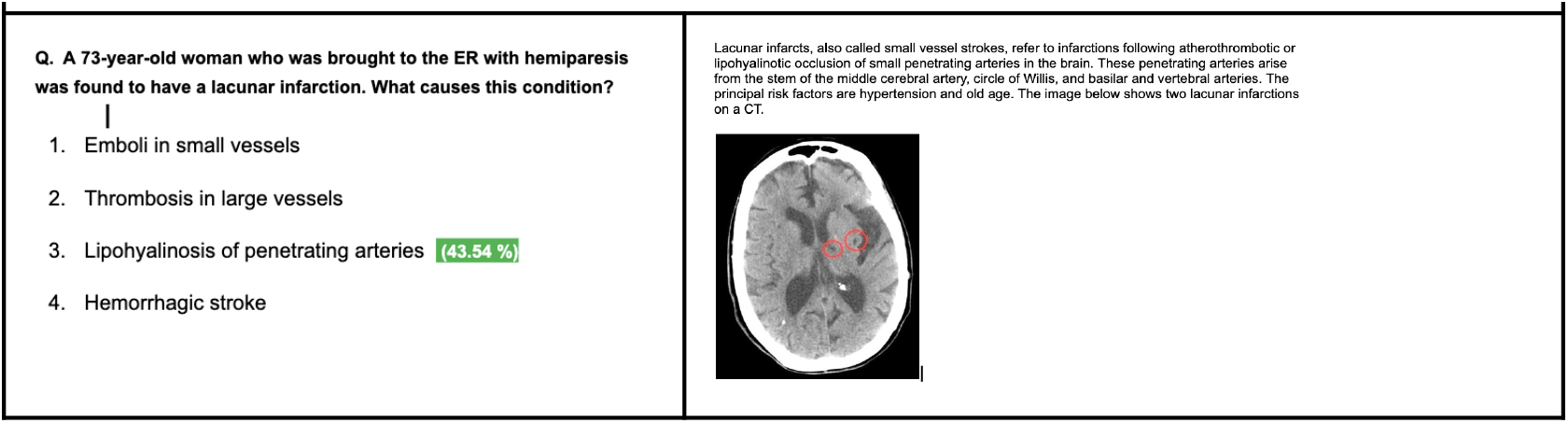
An sample MCQs from Schema topic: STEMI (ST Elevated Myocardial Infarction) Left side: Clinical scenario-based SOC-MCQ; Right side: Answer with explanation (shown after an attempt to solve); Note the percentage correctness in front of the correct option (For this particular MCQ, the total number of attempts = 282)

## Results

20 Schema topics were selected for the study comprising a total of 128 MCQs. The number of participants that solved each MCQ ranged from 60080 to 206672. A significant improvement in the net delta was observed in Anaphylaxis, STEMI, Covid-19 Vaccination, Glasgow Coma Scale, Subdural Hemorrhage, Syndromic Management of STIs, Laryngeal Mask Airway, and Basic Lifesaving Skills.

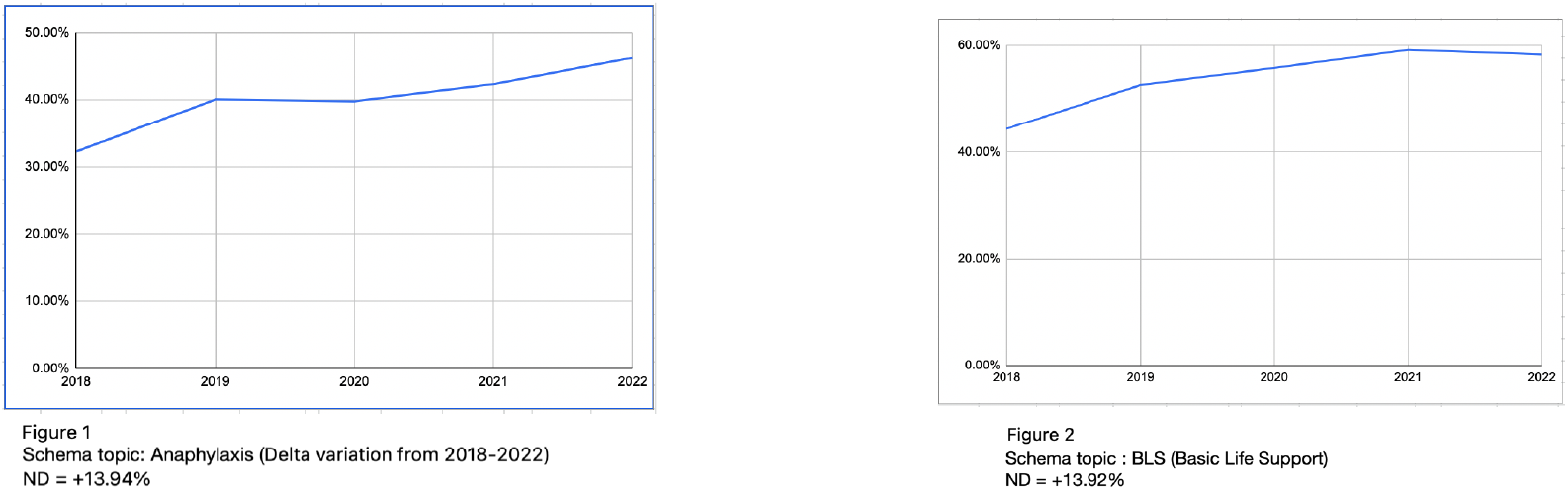

On the other hand, among the Schema topics that showed a significant (ND >5%) decline throughout 2018-2022 were Asthma management, Hypertension management, DKA (Diabetic Ketoacidosis), and SAH (Subarachnoid hemorrhage).

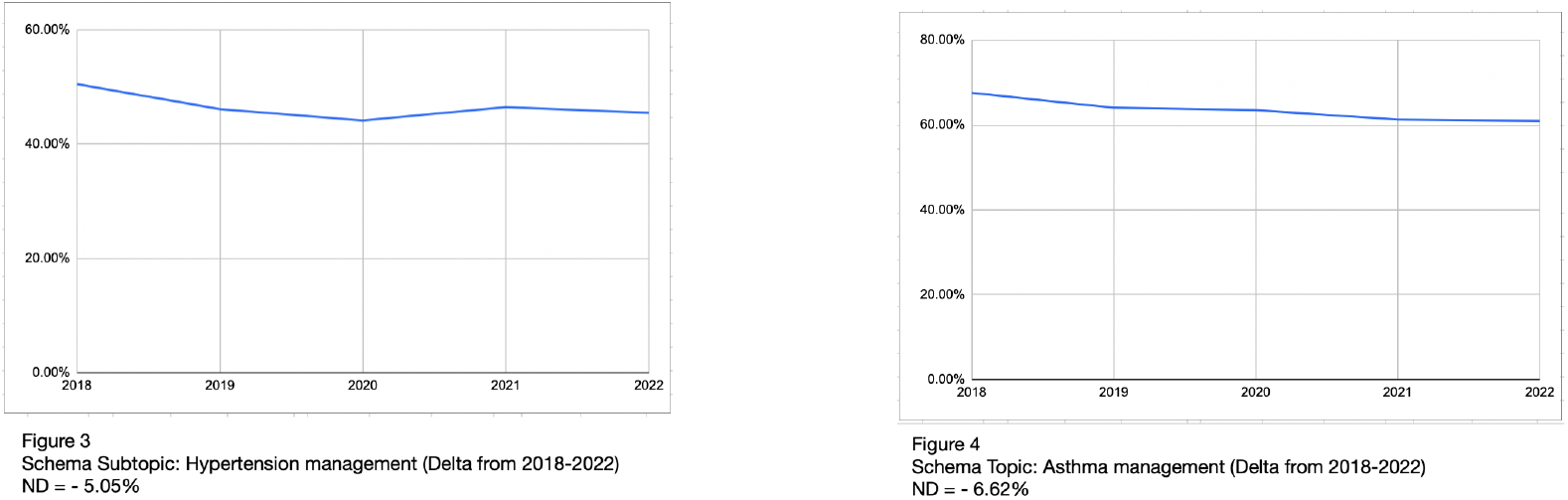

It is noteworthy that the Schema topic: *Covid-19 vaccination* is an exception to the inclusion criteria of MCQs attempted from 2018 to 2022 since it had been selected as a crucial topic but was only inculcated into Schema after 2019. Hence, the outlying ND of 60.52% is considered a consequence of this exception and not an anomaly.

**Table 2:** Cumulative observations of Net Delta (ND) of the selected Schema topics.

| Significant positive change<br>(ND ≥ 5%) | No significant change<br>(ND range: 5% to > -5% ) | Significant negative change<br>(ND ≤ 5%) |
| --- | --- | --- |
| Anaphylaxis (13.94 %)<br>STEMI (10.00 %)<br>Covid-19 Vaccination (60.52 %)<br>Glasgow Coma Scale (36.73 %)<br>Subdural Hemorrhage (5.29 %)<br>Syndromic Management of STIs (8.87 %)<br>Laryngeal Mask Airway (8.31 %)<br>BLS (13.97) | Ischemic Stroke - Management (1.40 %)<br>Ischemic Stroke - Risk Factors (2.46 %)<br>Vaccine Vial Monitoring (0.42 %)<br>Vaccine & Drug Storage (0.68 %)<br>Community-Acquired Pneumonia - Management (-4.96 %)<br>Extradural Hemorrhage (0.75 %)<br>NSTEMI (-3.96 %)<br>Acetaminophen (-1.57 %) | Asthma (-6.62 %)<br>Diabetic Ketoacidosis (-7.38 %)<br>Hypertension - Management (-5.05 %)<br>Subarachnoid Hemorrhage (-7.83) |

## Discussion

The complexity of medical education and practice presents unique challenges to applying quantified learning and assessment models. The intertwined web knowledge spread across segregated categories called ‘subjects’ also warrants deep horizontal and vertical integration, not just learning but also assessment of knowledge and competence. While many systems target conceptual learning & adaptiveness, the need for granulation of medical curriculum in terms of cruciality in applied medical practice remains an uncracked challenge. (32–34)

Schema not only makes such granulation and integration possible but also promotes a multidisciplinary assessment of knowledge & skill that is quantified and objective. It provides us with a metric that can be used to gauge the competence and potential of medical personnel. To the medical professionals themselves, it offers a reliable way to self-assess, acquire invaluable feedback, and, most importantly, the scope for improvement. In this study, 6 out of the 8 Schema topics that showed a significant uptrend in competence pertain to medical emergencies. It is an interesting correlation to note that recent trends in examination also show an uptrend in the number of questions in this category. Similarly, a downtrend was observed in Diabetes & Hypertension management.

As shown in this study, another application of Schema is the in-depth study and analysis of ever-changing trends & patterns in medical learning and education. For the first time, it is possible to acquire a microscopic or personal view of medical competence as well as a macroscopic or bird’s eye view of the medical fraternity as a whole. Such an application may be remarkably useful in medical education policy making, quality assurance interventions, and the successful deployment of learner-centric education programs.

The medical college of the future will amalgamate the imparting of medical education with innovative learning methodologies and disruptive technologies that will transform the currently existing education models. Artificial intelligence, data sciences, machine learning & quantified learning will be invaluable tools to influence learning effectively and foster academic growth.(35). Such technology not only enhances scopes of learning but also provides an alternative to the traditional learning models, helps in producing competent medical professionals, and increases the efficiency of learning & assessment so that the saved time can be used in pursuit of other ventures such as communicative development & social engagement.

### Future Scope of The Schema Method

The Schema Method puts holistic performance and skill enhancement at the center of learning. The continuous feedback to the learner combined with the extramural associations of topics across subjects undoubtedly contributes to a holistic understanding of Medicine.

It also provides us with a wide range of possible use cases in the future aimed toward performance analysis, knowledge gap redressal, learning pattern recognition, feedback effectiveness, comparative analysis of subjective vs. schema-based studies, and extensive knowledge maps based on Schema to explore the deeper interrelation of subjects.

Such uses are also likely to improve the understanding of the medical curriculum in the long run, which can be quantified using the Schema itself.

Assessment of medical students at the end of their training in terms of knowledge and practicing healthcare professionals in terms of patient care may also be a use case in the future.

The implementation of AI to develop a tailored approach to learning for each learner also falls well within the future scope of Schema.

**Table 3:**
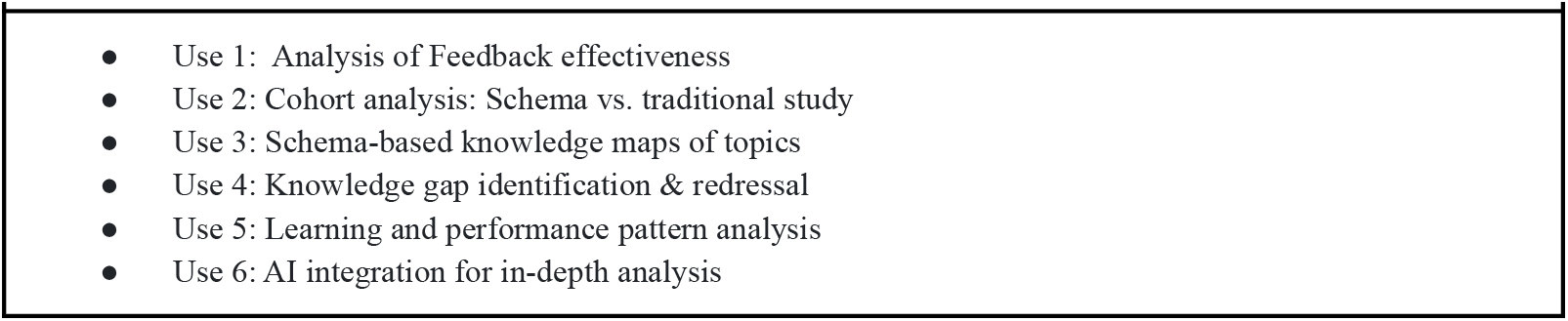
Future Use Cases of The Schema Method.

## Conclusion

This study explored Schema as a much-needed solution to the problem of the unavailability of quantified learning, assessment & feedback in the existing models of medical education. It also offers a mode to monitor the performances of millions of medical students and study the changing trends in medical education in real-time, so appropriate corrective steps can be taken by policymakers to reduce knowledge gaps wherever necessary. The future scopes of Schema make it an exciting tool for the further development of how we learn Medicine around the world.

### Limitations of Study

1. The study was done using retrospective user data from the database of a leading e-learning app for medical students. Consequently, the pool of participants changed throughout the study.
2. No blinding could be done as the study was retrospective.
3. It could not be ensured that all participants solved all MCQs in the study or that the same questions appeared to all participants.
4. It could not be assured that all participants were exposed to the same learning methodologies before the commencement of the study.
5. Most of the limitations mentioned above can be eliminated in future studies by opting for prospective study designs.

## Data Availability

All data produced in the present study are available upon reasonable request to the authors

